# Determinants of Health-Related Quality of Life (HrQoL) of People living with HIV Based on Sexual Orientation

**DOI:** 10.1101/2022.09.02.22279532

**Authors:** Rico Januar Sitorus, Nyoman Yudi Antara, Reymart V. Sangalang, Merry Natalia Panjaitan, Nelsensius Klau Fauk

## Abstract

**Purpose:** Human immunodeficiency virus (HIV) infection has detrimental impacts on the lives of different population groups living with HIV, including men who have sex with men (MSM). Using the World Health Organization Quality of Life Questionnaire (WHOQOL – HIV BREF), this study aimed to assess the Health-Related Quality of Life (HrQoL) of men living with HIV with different sexual orientations and determine the dominant influential factors.

**Methods:** This cross-sectional study involved 206 men living with HIV. They were recruited from Sriwijaya Plus Foundation and a medical facility providing antiretroviral therapy (ART). Data were analyzed using Chi-square and binomial logistic regression.

**Results:** The analysis showed that the percentage of MSM patients was more than non-MSM/heterosexual men, accounting for 68.9% of the total population. Furthermore, depression status, social stigma, family support, therapy duration, and suffering duration were significantly associated with the quality of life of MSM and non-MSM (p-value <0.001). The multivariate logistic regression revealed that the most dominant influential factor was depression status (PR=5.417; 95% CI=2.473-11.876), where the majority of the depressed patients were 5.417 times more at risk of lower life quality compared to others.

**Conclusion:** Depression can lead to low quality of life among HIV patients. The findings suggest that there is a need for the development of intervention programs that address the psychological and social needs of PLHIV or support them to cope with depression and the social stigma facing them. Support from families and health workers can help them cope with psychological and social challenges faced by PLHIV and increase community acceptance of PLHIV.

## Introduction

Human Immunodeficiency Virus (HIV) infection is one of the global public health issues with high morbidity and mortality rates.^1,2^ The UNAIDS data reported an estimated 37.7 million people living with HIV (PLHIV) globally in 2020, with Eastern and Southern Africa, and Asia and the Pacific at the top leading position, accounting for 20.6 million and 5.8 million PLHIV, respectively.^3^ In the context of Indonesia, the AIDS national report shows an estimation of more than five thousand PLHIV in 2021, with less than 50 percent (144,632) who are actively on antiretroviral therapy (ART).^4^

HIV infection has been reported as causing PLHIV a range of detrimental impacts including psychological, social (stigma and discrimination), and economic impacts.^5,6^ These impacts compounded with many other factors can also lead to further negative impacts and result in reduced or poor quality of life (QoL) of PLHIV. Studies investigating QoL of PLHIV have reported that PLHIV experienced poor QoL in several domains such as physical health, psychological health, level of independence, social relationship, environmental and spirituality domains.^7–9^

Studies have also suggested several factors associated with poor QoL of PLHIV in general, such as low level of educational attainment, unemployment status, perception of being ill, and dissatisfaction with sexual activity.^7,10–12^ The experiences of family food insecurity, polypharmacy use, the advanced state of HIV infection and psychiatric comorbidities are also factors associated with poor QoL among PLHIV.^11–13^ The use of illicit drugs which negatively influence the physical and mental well-being of PLHIV was also found to cause poor QoL among them.^14^ Stigma, perceived discrimination, adverse effects of ART, non-adherence to ART low family income are factors associated with poor QoL among men who have sex with men (MSM) living with HIV^15,16^.

Previous studies have also reported factors associated with good QoL of PLHIV, such as being employed, having no financial concerns, not having mental health issues (e.g., stress, depression, anxiety) and other medical comorbidities, higher education level, and undergoing ART.^8,10,14,17,18^ Other supporting factors for good QoL of PLHIV include the availability of social support, including emotional, tangible, and informational support, and having good coping strategies towards HIV-related difficult life situation.^19,20^ Similarly, studies with MSM living with HIV have reported that their better acceptance of the illness, family acceptance and peer support are positively associated with their better QoL.^21–24^

Despite a range of factors associated with poor or good QoL of PLHIV as reported in the aforementioned studies, none of these studies reported a comparison of QoL of MSM living with HIV and non-MSM living with HIV or heterosexual men globally. In the context of Indonesia, there is limited evidence on and studies exploring the QoL of different populations groups living with HIV.^21,23–25^ Therefore, this study aimed to compare the QoL of men living with HIV with different sexual orientations and determine the dominant factors that affect their QoL.

## Methods

### Participants and procedures

This study used a quantitative approach with a cross-sectional design, and the sample population consists of 1180 HIV patients. Subsequently, a total of 206 respondents (MSM= 142; non-MSM=64) were selected using a non-random sampling technique, namely purposive sampling. They were registered in the Care Support and Treatment (CST) service and the Sriwijaya Plus Community in Palembang City The respondents’ QoL was measured using the World Health Organization Quality of Life-HIV BREF (WHOQOL-HIV BREF) instrument, which consisted of 6 domains, namely physical, psychological, level of independence, social relationships, environment, and spirituality. Each domain was rated on a 5-point Linkert scale where 1 indicates low and negative perceptions, while 5 shows high positive perceptions. The validity and reliability tests of the instrument were carried out in Indonesian. The validity test revealed a strong correlation coefficient (r = 0.60 – 0.79), while the Cronbach Alpha value obtained from the reliability test was in the medium and good categories (0.513-0.798).^26^

#### Depressive symptoms

The symptoms include the psychological state of the respondents within the last two weeks. Furthermore, their depression status was measured using the Patient Health Questionnaire-9 (PHQ-9), and the answers were given different scores, namely never (0), several days (1), more than a week (2), and almost every day (3). The PHQ was a self-administered version of the PRIME-MD diagnostic instrument for common mental disorders. The depression status was grouped into 2 categories, where scores of 5-27 indicated a depressed state, while 0-4 were categorized as not depressed.^27^

#### Social stigma

This is a bad mark or view received by PLWHA, and it was measured using the Berger HIV Stigma Scale instrument, where the total score ranged from 25-125. The categorization was carried out using a cut-off point formula of 75% of the total score (125), where values ≥ 93.75 indicated high stigma, while values < 93.75 were in the low category. The validity and reliability test of the Indonesian version of the Berger HIV Stigma Scale questionnaire (40 items) conducted by Nurdin obtained a Cronbach Alpha value of 0.94. Meanwhile, a value of 0.93 was recorded from the short version, which consisted of 25 items.^28^

#### Family support

This includes the support received by PLHIV from family members, such as husband, wife, and children as well as biological father, mother, brother, and sisters that cared for them during illness. It can be in various forms, including informational, emotional, instrumental, and appreciative support, which were measured using an instrument developed by Arikunto (2002), where the total score ranged from 18 to 90. The categorization was carried out using a cut-off point formula of 75% of the total score, where values < 67.5 indicate low support, while others ≥ 67.5 were in the high category. A Cronbach’s Alpha value of 0.6 was obtained from the reliability results.^29^

#### Occupation

Information about the respondent’s occupation was obtained through interviews with questions that were already available in a structured questionnaire. Their occupations were then categorized into “not working and working” for further analysis.

##### Duration of ART

Information about the respondent’s duration of ART was obtained through interviews using questions that were already available in a structured questionnaire. The durations were categorized into “< 1 year and ≥ 1 year” for further analysis.

#### Duration of living with HIV

Information about the respondent’s duration of HIV infection was collected through interviews with questions that were already available in a structured questionnaire. The durations were then categorized into “< 5 years and ≥ 5 years” for further analysis.

### Statistical analysis

Data were analyzed statistically in 3 stages. A univariate analysis was carried out to describe the characteristics of the respondents, after which the Chi-square test was applied to explore the relationship between the main independent variables and QoL. To determine the dominant factors, a multivariate analysis was performed using multiple logistic regression test.

## Results

Based on descriptive analysis, the demographic characteristics of the respondents showed that the percentage of MSM patients was more than non-MSM/heterosexual men, accounting for 68.9% of the total population. Table 1 revealed that within the MSM group, 68.1% had a job, 71.2% had incomes below the minimum wage, 71.2% were not married, and 76.9% were below the age of 30 years. Furthermore, 64.8% had an undergraduate degree, 68.8% were undergoing ART, and 70.2% were not depressed. The analysis also showed that 63.9% of the MSM participants experienced stigma, 72% had been diagnosed with HIV for less than 5 years, 80.6% received low family support, and 71% had a low QoL.

**Table 1.** Demographic Characteristics PLWHA based on sexual orientation

| Variables | Sexual orientation |  |
| --- | --- | --- |
|  | MSM<br>N=142 | Non-MSM<br>N=64 |
| Gender |  |  |
| Male | 142 (68,9 %) | 64 (31,1 %) |
| Occupation |  |  |
| - Unemployed | 18 (75,0 %) | 6 (25,0 %) |
| - Employed | 124 (68,1 %) | 58 (31,9 %) |
| Marital status |  |  |
| - Not married yet | 126 (82,9 %) | 26 (17,1 %) |
| - Married | 12 (27,3 %) | 32 (72,7 %) |
| - Widower | 4 (40,0 %) | 6 (60,0 %) |
| Income |  |  |
| - < regional minimum wage | 94 (71,2 %) | 38 (28,8 %) |
| - ≥ regional minimum wage | 48 (64,9 %) | 26 (35,1 %) |
| Age (years) |  |  |
| - < 30 | 80 (76,9 %) | 24 (23,1 %) |
| - ≥ 30 | 62 (60,8 %) | 40 (39,2 %) |
| Degree |  |  |
| - Elementary school | 2 (28,6 %) | 5 (71,4 %) |
| - Junior High School | 7 (70,0 %) | 3 (30,0 %) |
| - Senior High School | 81 (72,3 %) | 31 (27,7 %) |
| - Diploma | 15 (75,0 %) | 5 (25,0 %) |
| - Undergraduate | 35 (64,8 %) | 19 (35,2 %) |
| - Graduate | 2 (66,7 %) | 1 (33,3 %) |
| Quality of Life |  |  |
| - Low | 76 (71,0 %) | 31 (29,0 %) |
| - High | 66 (66,7 %) | 33 (33,3 %) |
| Social stigma |  |  |
| - High | 43(84,3 %) | 8 (15,7 %) |
| - Low | 99 (63,9 %) | 56 (36,1 %) |
| Duration of ARV therapy (years) |  |  |
| - < 1 | 23 (69,7 %) | 10 (30,3 %) |
| - ≥ 1 | 119 (68,8 %) | 54 (31,2 %) |
| Duration of living with HIV (years) |  |  |
| - < 5 | 118 (72,0 %) | 46 (28,0 %) |
| - ≥ 5 | 24 (57,1 %) | 18 (42,9 %) |
| Family support |  |  |
| - Low support | 108 (80,6 %) | 26 (19,4 %) |
| - High support | 34 (47,2 %) | 38 (52,8 %) |
| Depression symptoms |  |  |
| - Depression | 36 (65,5 %) | 19 (34,5 %) |
| - Not depression | 106 (70,2 %) | 45 (29,8 %) |

The evaluation of the participants’ QoL showed that non-MSM/heterosexual men had a better QoL in the physical domain. However, the MSM group was better in the psychological aspect, independencies, social interaction, environmental domain, and perception of health, as shown in Table 2. The bivariate analysis showed that depression status, social stigma, family support and the duration of ART and HIV infection significantly correlated with the QoL of MSM and non-MSM with a p-value of 0.000, as shown in Table 3.

**Table 2.**
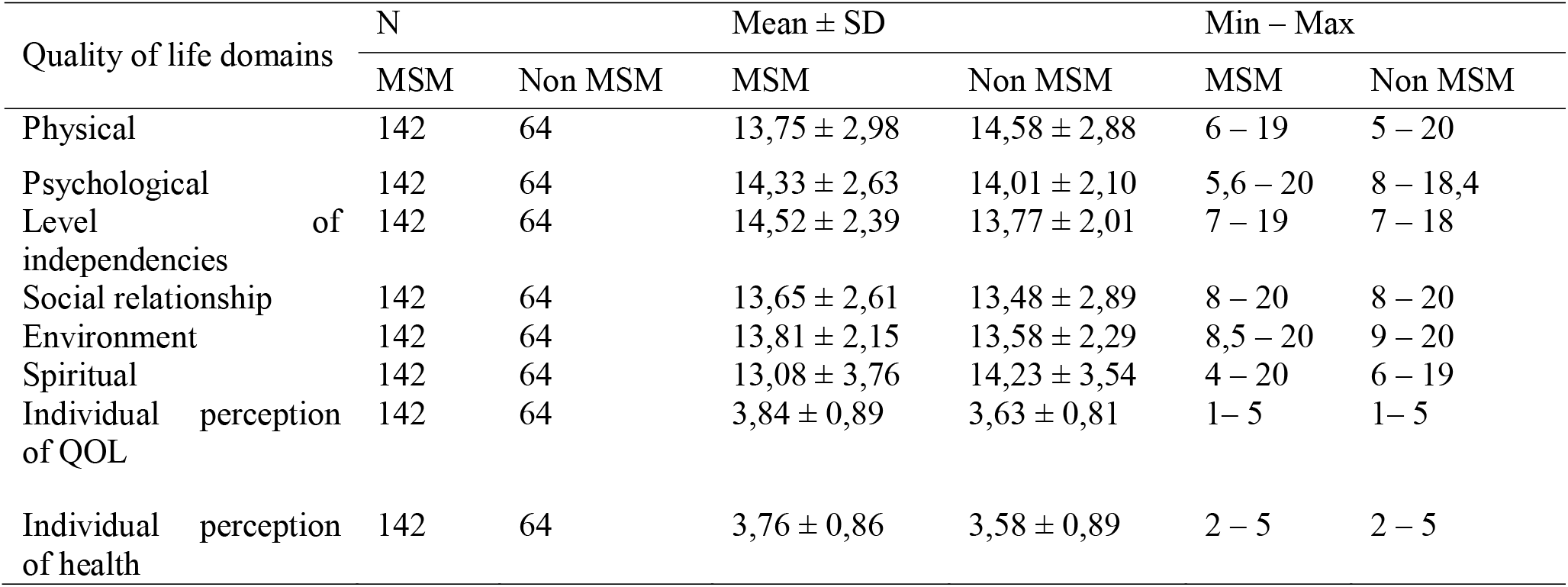
Descriptive analysis Health-Related Quality of Life (HrQoL) of PLWHA based on sexual orientation

| Quality of life domains | N | | Mean ± SD | | Min – Max | |
| --- | --- | --- | --- | --- | --- | --- |
|  | MSM | Non MSM | MSM | Non MSM | MSM | Non MSM |
| Physical | 142 | 64 | 13,75 ± 2,98 | 14,58 ± 2,88 | 6 – 19 | 5 – 20 |
| Psychological | 142 | 64 | 14,33 ± 2,63 | 14,01 ± 2,10 | 5,6 – 20 | 8 – 18,4 |
| Level of independencies | 142 | 64 | 14,52 ± 2,39 | 13,77 ± 2,01 | 7 – 19 | 7 – 18 |
| Social relationship | 142 | 64 | 13,65 ± 2,61 | 13,48 ± 2,89 | 8 – 20 | 8 – 20 |
| Environment | 142 | 64 | 13,81 ± 2,15 | 13,58 ± 2,29 | 8,5 – 20 | 9 – 20 |
| Spiritual | 142 | 64 | 13,08 ± 3,76 | 14,23 ± 3,54 | 4 – 20 | 6 – 19 |
| Individual perception of QOL | 142 | 64 | 3,84 ± 0,89 | 3,63 ± 0,81 | 1 – 5 | 1 – 5 |
| Individual perception of health | 142 | 64 | 3,76 ± 0,86 | 3,58 ± 0,89 | 2 – 5 | 2 – 5 |

**Table 3.** Predictors of Health-Related Quality of Life (HrQoL)

| Variable | Category | Quality of life |  |  |  | p-value | PR (95% CI) |
| --- | --- | --- | --- | --- | --- | --- | --- |
|  |  | Low |  | High |  |  |  |
|  |  | n | % | n | % |  |  |
| Depression status | Depression | 45 | 81.8 | 10 | 18.2 | 0,000 | 1,993 |
|  | Not depression | 62 | 41.1 | 89 | 58.9 |  | (1,586 – 2,503) |
| Social stigma | High | 36 | 70.6 | 15 | 29.4 | 0,004 | 1,541 |
|  | Low | 71 | 45.8 | 84 | 54.2 |  | (1,204 – 1,972) |
| Family support | Low | 80 | 59.7 | 54 | 40.3 | 0,004 | 1,592 |
|  | High | 27 | 37.5 | 45 | 62.5 |  | (1,146 – 2,212) |
| Occupation | Unemployed | 16 | 66.7 | 8 | 33.3 | 0,187 | 1,333 |
|  | Employed | 91 | 50.0 | 91 | 50.0 |  | (0,970 – 1,833) |
| Duration of ARV Therapy | < 1 year | 23 | 69.7 | 10 | 30.3 | 0,042 | 1,435 |
|  | ≥ 1 year | 84 | 48.6 | 89 | 51.4 |  | (1,093 – 1,885) |
| Duration of living with HIV | < 5 years | 91 | 55.5 | 73 | 44.5 | 0,066 | 1,457 |
|  | ≥ 5 years | 16 | 38.1 | 26 | 61.9 |  | (0,967 – 2,193) |

The multivariate analysis using logistic regression revealed that the most influential factor was depression status (p-value=0.000, PR _Adj_ 5.417; 95 % CI (2.473-11.867) in both groups MSM and non-MSM. The finding indicates that depressed HIV patients were 5.417 times more at risk than others, as shown in Table 4.

**Table 4.**
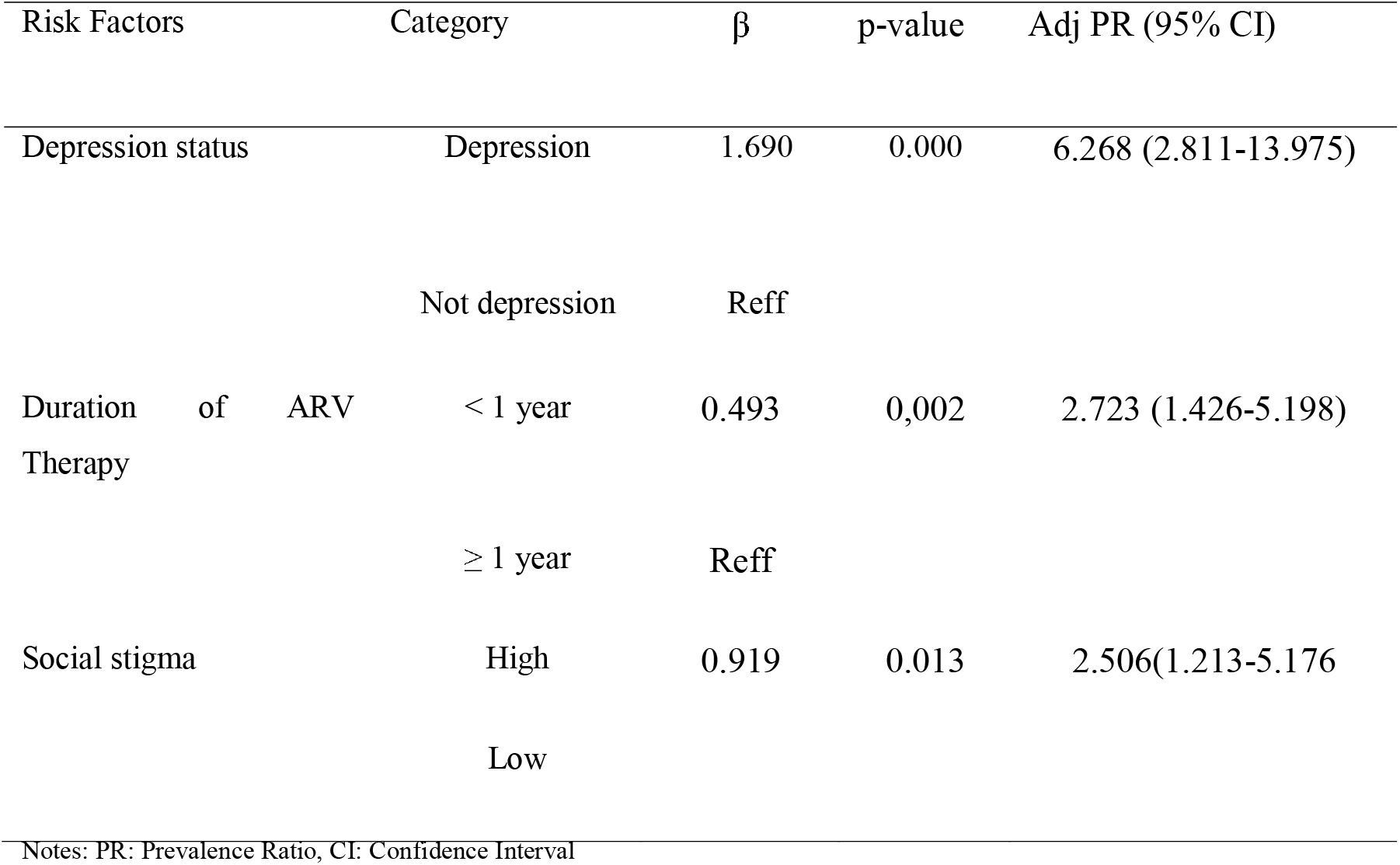
Multivariate analysis of Health-Related Quality of Life (HrQoL)

| Risk Factors | Category | $\beta$ | p-value | Adj PR (95% CI) |
| --- | --- | --- | --- | --- |
| Depression status | Depression | 1.690 | 0.000 | 6.268 (2.811-13.975) |
|  | Not depression | Reff |  |  |
| Duration of ARV Therapy | < 1 year | 0.493 | 0,002 | 2.723 (1.426-5.198) |
| | $\geq 1$ year | Reff | | |
| Social stigma | High | 0.919 | 0.013 | 2.506(1.213-5.176) |
|  | Low |  |  |  |

Notes: PR: Prevalence Ratio, CI: Confidence Interval

## Discussion

The study suggests that the MSM group had lower QoL compared to the non-MSM/ heterosexual men. The higher prevalence rate of depression among the MSM group compared to the non-MSM group was a possible explanation for their experience of low QoL. Our findings confirm previous findings which have reported that depression is common among PLHIV due to their experience of health status deterioration, negative side effects of ART and apathy following their HIV diagnosis.^30–33^

In addition, some study suggests that depression, which consists of a series of disorders, can have negative effects on sleep, weight loss, appetite, health-seeking behaviors, and motivation of PLHIV, which in turn can further deteriorate their health and psychological wellbeing.^34^ It has also been reported that depression is one of the most common psychiatric disorders, which negatively impacts the adherence and outcomes of ART among PLHIV.^35,36^

Our findings show that perceived and internalized stigma led to an increase in the intensity of depression among MSM and non-MSM. They also influenced both groups of patients’ health behaviors and led to non-disclosure of HIV status to partners, poor adherence to ART, increased risk of developing drug resistance, restricted access to health services, and reduced HrQoL. These are in line with the results of previous studies suggesting the association of stigma with poor physical life quality among PLHIV.^37–40^

It has been well-documented that despite numerous efforts to reduce the negative impact of HIV-related stigma, patients or PLHIV are still stigmatized in various contexts, including within families, communities, workplaces, and healthcare settings.^38,41–43^ This study also suggests that there is a significant relationship between QoL and family support in both MSM and non-MSM with HIV. The finding is consistent with the findings of previous studies reporting that family support is associated with encouragement and the absence of stigma and discrimination against PLHIV.^44^

Strategies to improve the quality of life of PLHIV are by strengthening support and family care for them and promoting HIV screening among high-risk populations. Family supports include informing medication adherence, overcoming discrimination, encouraging early ART initiation, and attending therapy regularly can reduce loss to follow-up.^45^

Adherence to therapy strongly supports the QoL of PLHIV. This study indicates that the length of treatment is significantly related to improving the quality of life of PLHIV. This is consistent with the findings of previous studies which have suggested that being on ART is a factor associated with good QoL of PLHIV.^8,10^

The intake of ART helps to lower viral load, improve physical immune function and reduce opportunistic infections and comorbidities. It also increases the patient’s productivity, social inclination, and QoL.^46^ ART has also become a key component that increases longevity and controls other infectious diseases and has a significant long-term contribution to improving HrQoL.^47,48^

## Conclusion

The results showed that the average physical and spiritual life quality of HIV patients with MSM sexual orientation was higher than others in the non-MSM category. They also had better psychological well-being, independence level, social relationships, environment, and perceptions of health. The patient’s life quality can be improved by providing vital support, reducing stigma as well as paying attention to stress levels and therapy adherence.

## Data Availability

All data produced in the present work are contained in the manuscript

## Acknowledgment

The authors are grateful to the manager of Mohammad Hoesin Hospital as well as the medical record unit staff. The authors are also grateful to Sriwijaya Plus Foundation, Dempo community health center, and Sukarami community health center.

## Authors’ Contribution

R.J.S. designed the study, developed a data instrument for data collection analysis, and drafted the manuscript. N.K.F. contributed to the proofreading and drafted the manuscript. N.Y.A. contributed to the interpretation of results, as well as the reviewing and editing of the article. Furthermore, R.V.S. contributed to the proofreading and editing of the article, while M.N.P. assisted in the literature review and editing. All co-authors reviewed and approved the final manuscript before submission.

## Ethics Approval and Consent to Participate

This study was approved by the ethical review committee of the Faculty of Public Health Sriwijaya University with reference number 149/UN9.FKM/TU.KKE/2021.

## Competing Interest

The authors declare no significant competing financial, professional, or personal interests that can affect the performance or presentation of the work described in this manuscript.

## Availability of Data and Materials

All data and related materials from this study are available and can be provided by the first author

## Notes

### Competing Interest Statement

The authors have declared no competing interest.

### Funding Statement

This study did not receive any funding

